# Analysis of whole exome sequencing in severe mental illness hints at selection of brain development and immune related genes

**DOI:** 10.1101/2021.05.11.21257013

**Authors:** Jayant Mahadevan, Ajai Kumar Pathak, Alekhya Vemula, Ravi Kumar Nadella, Biju Viswanath, Meera Purushottam, Sanjeev Jain, Mayukh Mondal

## Abstract

Evolutionary trends may underlie some aspects of the risk for common, non-communicable disorders, including psychiatric disease. We analyzed whole exome sequencing data from 80 unique individuals from India coming from families with two or more individuals with severe mental illness. We used Population Branch Statistics (PBS) to identify variants and genes under positive selection and identified 75 genes as candidates for positive selection. Of these, 20 were previously associated with Schizophrenia, Alzheimer’s disease and cognitive abilities in genome wide association studies. We then checked whether any of these 75 genes were involved in common biological pathways or related to specific cellular or molecular functions. We found that immune related pathways and functions related to innate immunity such as antigen binding were over-represented. We also evaluated for the presence of Neanderthal introgressed segments in these genes and found Neanderthal introgression in a single gene out of the 75 candidate genes. However, the introgression pattern indicates the region is unlikely to be the source for selection. Our findings hint at how selection pressures in individuals from families with a history of severe mental illness may diverge from the general population. Further, it also provides insights into the genetic architecture of severe mental illness, such as schizophrenia and its link to immune factors.

## Introduction

Severe mental illnesses (SMI) such as schizophrenia and bipolar disorder (BD) have a lifetime prevalence of 1%; and this seems to have remained relatively stable across geography and time ^1,2^. Psychiatric disease syndromes are common, usually begin in young adulthood, are a source of considerable personal and social distress, associated with premature mortality and the treatments have limited efficacy. Hence, detecting underlying mechanisms that may contribute to risk and recovery will be useful.

These syndromes are known to be heritable and their genetic architecture is quite likely to be polygenic, with a combination of common variants of small effect and rare variants of relatively larger effect being implicated ^3^. The apparently uniform genetic epidemiology of these syndromes in different parts of the world seems to suggest that there is no specific selection for, or against, these conditions. This has been attributed to theories of balancing selection, ancestral neutrality or polygenic mutation selection balance ^4^.

From a population genetics standpoint, admixture, migrations and selection all have an impact on our understanding of the genetic contributions to risk of mental illness^5^. Genome wide association study (GWAS) data and summary statistics have been most commonly used to investigate the contribution of natural selection on the genetic architecture of complex traits, such as psychiatric syndromes ^6^. Findings from studies in mental illness have been ambiguous with a few implicating the role of positive selection ^7–9^, while a number of others have shown either no evidence for selection or negative selection ^4,10,11^.

Whole exome sequencing, which documents the variation in protein-coding sequences, has also been used as a tool to investigate natural selection. A number of studies in isolated populations have identified genetic variation that confers protection against environmental conditions such as adaptation to hypoxia at high altitudes among Tibetans ^12^ or arctic climate among Nunavik Inuit ^13^ and Siberians ^14^. Signatures of natural selection are detected even in the context of more recent population divergence and this influences many aspects of physiology, underscored by variations in genes that impact on height, blood coagulation, pigmentation, diet availability and resistance to infections ^15,16^.

In addition to the role of natural selection, there has also been growing interest in understanding the contribution of archaic sequences of DNA to liability for disease ^17^. We know that there has been more than one instance of admixture between early human populations along with Neanderthals and Denisovans ^18,19^. This has resulted in the persistence of a number of introgressed sequences of archaic (Neanderthal and Denisovan) DNA that account for around 2 - 4% of the genome in modern (*Homo sapiens*) human populations. Studies have demonstrated the influence of such sequences on immune functioning and susceptibility to infections including COVID - 19 ^20^. These sequences have also been found to be depleted in genes related to specific brain regions ^21^ and influence functional connectivity in the brain as well ^22^. Consequently, the impact of archaic introgression and admixtures on psychiatric disorders merits further exploration.

South Asia has been inhabited by modern humans for the last several thousand years, and the population displays admixture with both extinct hominins, as well as significant migrations and bottlenecks in the recent past ^23,24^. These admixture events may thus have a noteworthy influence on the susceptibility and prevalence of disease.

Hence, in this study, we investigated signatures of selection in unrelated individuals from families with multiple affected individuals with severe mental illness from southern India. We also used allele frequency differences between the cases and controls from the same population to prune out potential regions directly associated with caseness, and concentrated on regions with strong positive selection. Additionally, we specifically explored whether genes which showed evidence of selection had any evidence of Neanderthal introgression.

## Results

We used whole exome sequence (WES) data of 80 unique individuals from families in which multiple members (at least 2 first-degree relatives in a nuclear family) were diagnosed to have a major psychiatric disorder as cases for the analysis ^25^. Since WES data is highly dispersed, we decided to use Population Branch Statistics (PBS) for our selection analysis ^26^. PBS is based on allele frequency differentiation between populations using three populations. Unlike F_ST_ ^27^, PBS is directional and gives us a clear idea as to which population is under selection for the given allele. High PBS value corresponds to a highly deviated allele frequency of the target population compared to the reference population caused by positive selection. Here, we used our data set consisting of 80 cases as the target population, the South Asian and African ancestry genomes from the GnomAD dataset as reference and outgroup populations respectively ^28^. We also used WES data from 10 individuals as controls to exclude PBS differences that may be attributable to case status rather than selection (See below for more details).

### Identification of SNPs and genes under influence of selection

We followed an approach that defined the genes that fell in the 99.9th percentile of the PBS value distribution as the most likely candidates for selection. Further, to increase the confidence that the SNPs that fell under extreme PBS values were caused by selection rather than sampling of cases, we calculated the frequency difference of these SNPs between cases and controls and chose only those SNPs that were not in top 99.9 percent of the frequency difference. This provided a list of candidate genes which were a plausible target of adaptive evolution specific to our test population. Further, we filtered genes with at least two SNPs with high PBS value to reduce the chance of false positives. A total of 398 SNPs located in 185 genes were found to lie in the top 0.1 % of the PBS. Of these, 110 genes had only one SNP per gene (Supplementary Table S1), while 75 genes out of these 185 genes had more than one SNP per gene (Supplementary Table S2). For these 75 genes, we calculated an average PBS value per gene (Figure 1; Supplementary Table S3). We then used this list of 75 genes to look for any indications of an underlying shared biology.

**Figure 1:**
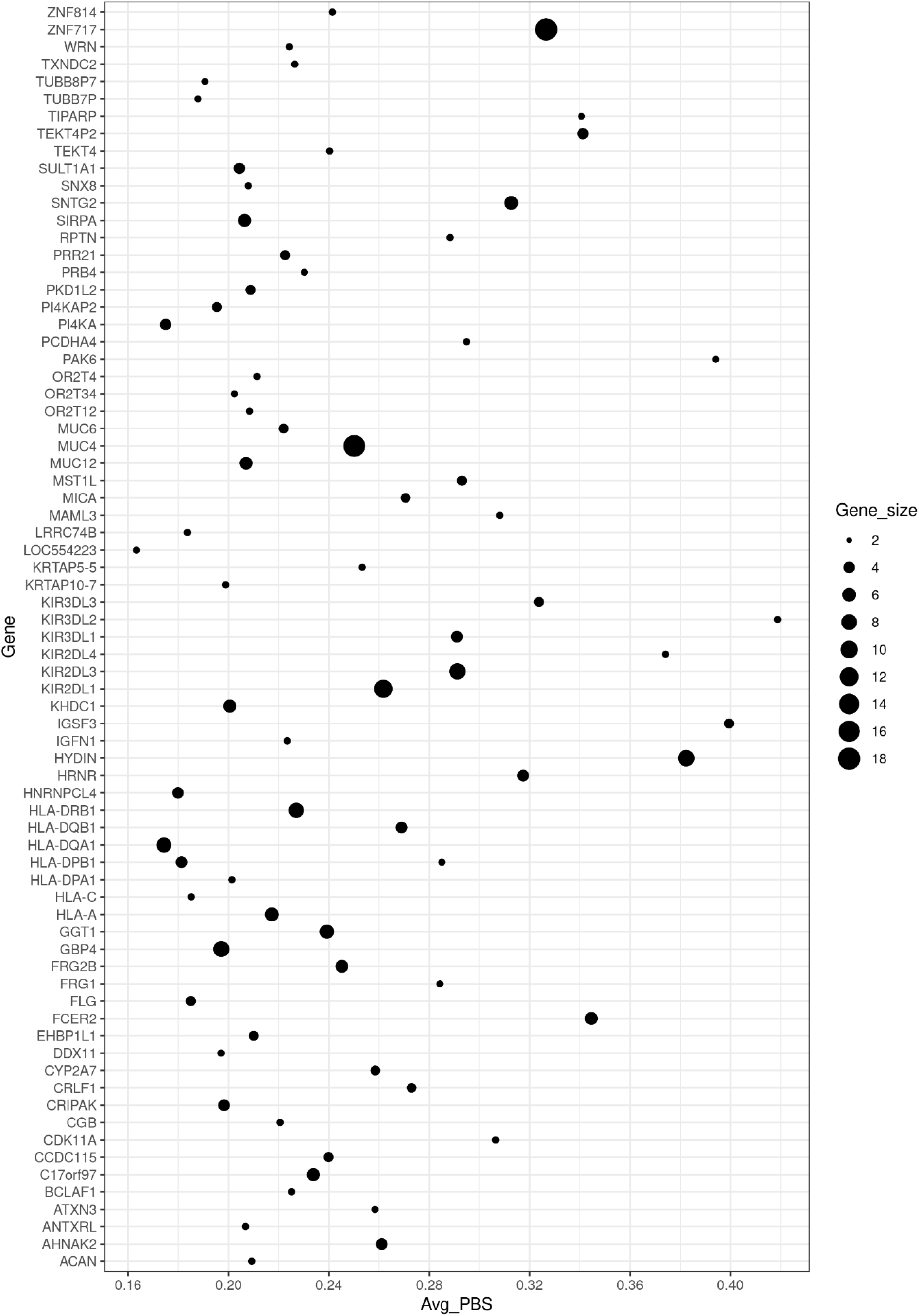
Dot plot of genes with multiple SNPs against the average PBS value; the dot size varies based on gene size (number of SNPs). While plotting we removed one of the gene -SARM1- that was behaving as an outlier (average PBS value = 0.89591), for better visualization.

### Functions of putatively selected genes

Many of the 75 genes are involved with immunological and defense responses including activation and regulation of interferon-gamma, cytokine and immune system, and different signaling pathways.

We manually curated these genes using the GWAS Catalogue (https://www.ebi.ac.uk/gwas/) and ENSEMBLE (https://www.ensembl.org/Homo_sapiens/Info/Index) and found that several of the listed genes were reported for having potential roles in cancer, liver disease and diabetes.

We found 20 genes (MAML3, SNX8, WRN, ATXN3, PAK6, TXNDC2, MICA, PK1LD2, AHNAK2, PI4KA, C17orf97, FCER2, SNTG2, GGT1, FLG, IGFN1, PCDHA4, ANTXRL SULT1A1), in this list of 75 genes, that have been previously associated with elevated risk for schizophrenia, Parkinson disease, Alzheimer’s Disease and cognitive abilities or intelligence (Supplementary Table S4).

### Functional enrichment and pathway enrichment analysis

Furthermore, to evaluate whether genes with extremely high PBS values (the 99.9th percentile) were enriched in any functional category or metabolic pathways, we evaluated our list of 75 genes for the three Gene Ontology (GO) categories: biological processes (Supplementary Table S5), cellular components (Supplementary Table S6), and molecular function (Supplementary Table S7). In addition, we analyzed the gene list for pathway over-representation using the IMPaLa tool (Supplementary Table S8).

In GO analysis, we observed that several of selected genes of the target population were functionally enriched (P < 0.05) for different signaling and regulatory mechanisms related to immune system, and viral defense such as negative regulation of natural killer cell mediated cytotoxicity, interferon-gamma-mediated signaling pathway and antigen processing and presentation of exogenous peptide or polysaccharide antigen via MHC class II.

Using the IMPaLa pathway analysis tool, we again observed an over-representation for the enrichment of genes (below Q-value 0.05) involved in different immune related pathways including antigen processing and presentation, Graft-versus-host disease, Type I diabetes mellitus and autoimmune thyroid disease.

We repeated the functional enrichment analysis and pathway analysis with our list of 75 genes after exclusion of the HLA region (which is known to be extremely polymorphic). It was seen that while the GO analysis did not show any evidence of functional enrichment, the pathway analysis using IMPaLA suggested pathways involved with immune function (Table1).

**Table 1:**
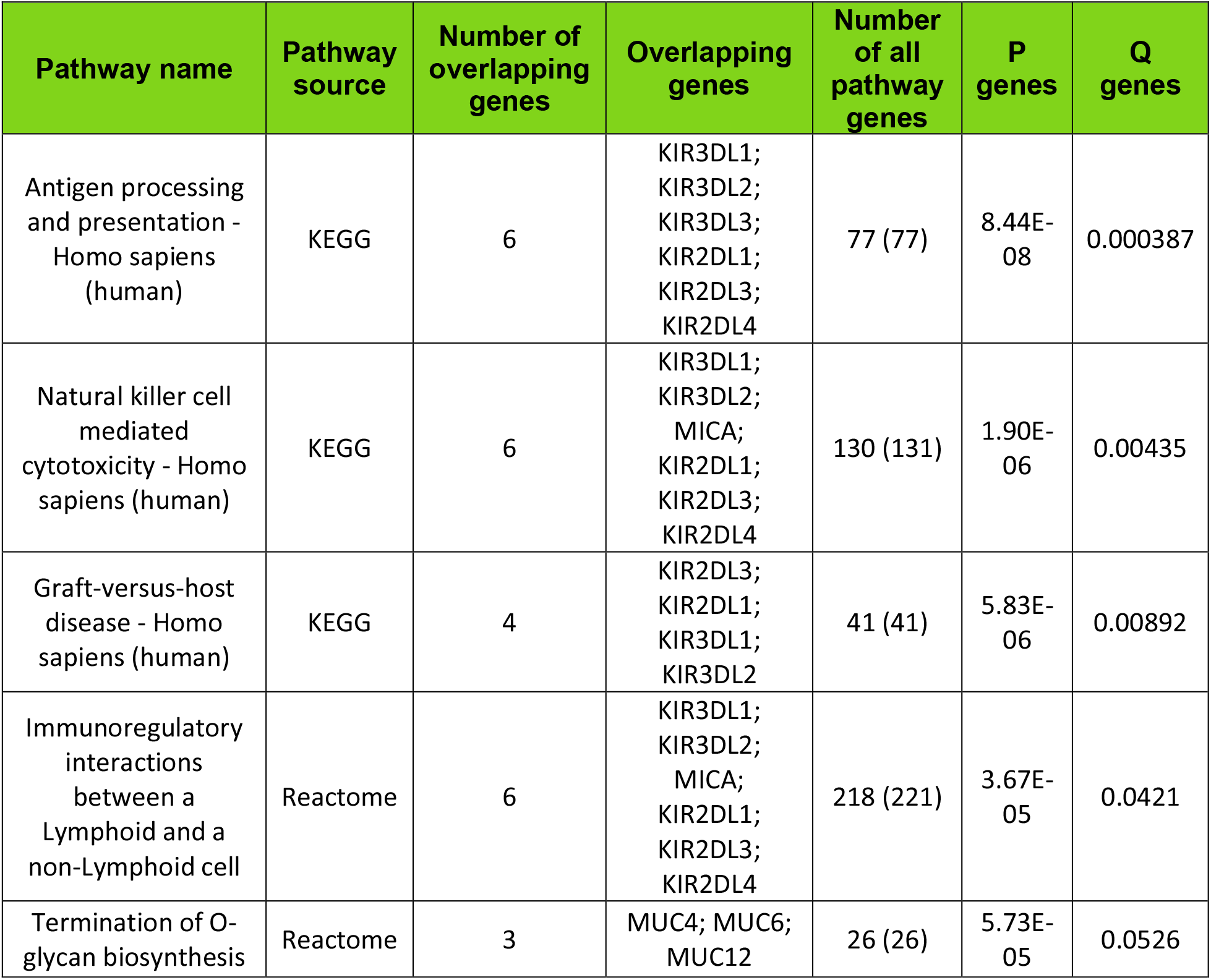
IMPaLa pathways enrichment analysis results for the list of 75 multiSNP genes after removing HLA genes.

### Archaic introgression in putatively selected genes

Further, we looked for the archaic introgression in these 75 genes that were chosen as candidates of selection. We applied Haplostrips ^29^ to the exome data of our 80 unrelated case samples. However, we could not detect any definitive traces of archaic introgression in any of the genes in the gene sets except *AHNAK2* gene (Figure 2). Though we observed a few Neanderthal derived SNPs in the AHNAK2 gene region showing a pattern of unique haplotype sharing among the continental populations, none of these SNPs were found in high PBS value during selection scan.

**Figure 2:**
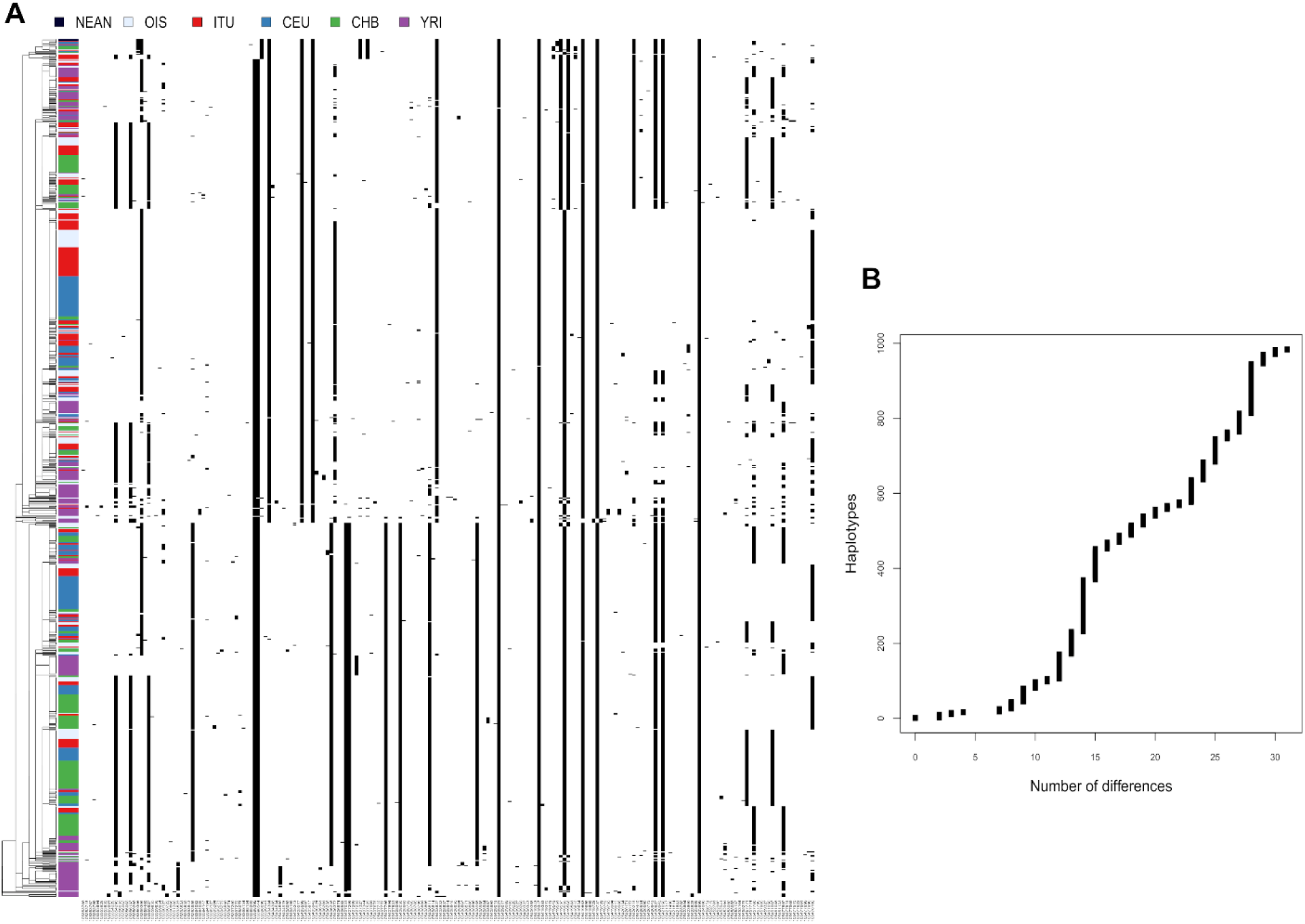
Haplostrips plot of AHNAK2: (A) clustered and sorted by increasing distance with Neanderthals. a few Neanderthal derived SNPs show a pattern of unique haplotype sharing among the continental populations, however, none of these SNPs were found in high PBS value during selection scan. Therefore, mostly have no significance in adaptive role of AHNAK2 in this study particularly; population label abbreviations are as follows: NEAN, Neadnrthal; OIS, Our Indian Samples; ITU, Indian Telugus; CEU, Central Europeans from Utah; CHB, Chinese Han from Beijing; YRI, Yoruba. (B) this plot visualizes the extent of closeness (based on SNP difference) between the haplotypes shared by continental populations and Neanderthal.

The fact that Neanderthal introgression sequence in the *AHNAK2* gene region does not contain any of the variants identified as putative candidates of selection, indicates that the introgressed haplotype of the gene was probably not selected, thus, rules out its significance in the adaptive role of AHNAK2 in this study.

## Discussion

Our results identify a total of 75 genes that show definitive evidence of selection, in individuals coming from families with multiple individuals with severe mental illness from southern India, based on the PBS analysis. Many of the identified genes have complex biology with many being linked to immune processes, cancer and neuropsychiatric illnesses.

There is some prior suggestion that many genes implicated in brain function, and disease, may be under selection ^30^. This was supported by our findings where genes that we identified to be under selection such as *SNX8* and *PAK6*, were seen to harbour common variants which have been associated with the risk for schizophrenia in GWAS ^10,31–33^. Genes identified such as *SNTG2* and *PKD1L2* were associated with other neurodevelopmental phenotypes such as autism ^34^ and attention deficit hyperactivity disorder ^35^. We also found that genes related to disorders of neuronal migration and brain malformations such as polymicrogyria, like *AHNAK2* and *PI4KA*; and genes harbouring variants linked to intelligence such as *MAML3, WRN* and *SULT1A1* were under selection ^36–38^. At the other end of the spectrum, some of the genes we identified such as *ATXN3* and *C17orf97* have also been known to harbour variants associated with neurodegenerative disorders like Amyotrophic Lateral Sclerosis ^39^ and Parkinson’s disease ^40^.

A number of identified genes have also been implicated in brain biology, including neurodevelopment, maintenance of neuronal and axonal integrity and apoptosis. The SARM1 protein is a member of MyD88/TIR-domain containing superfamily of proteins, which are involved in innate immune responses. Deficiency of the protein is seen to influence the apoptotic cascade of a variety of neural cells, including microglia; and cytokine expression in the brain ^41^. The HYDIN protein, associated with cilia in the brain, are critical for development of ventricles and brain; and it interacts with other genes like *FOXP2* which are well known to be related to neurodevelopmental disorders such as ASD and ADHD ^42^. The neurodevelopmental disorders of autism and schizophrenia have also been hypothesised to be linked to UV exposure possibly via vitamin D levels ^43^. Thus, it was interesting to identify the *HORNERIN / HRNR* gene which is very sensitive to UV light and is believed to also be involved in the species-specific differentiation of the outer layer of skin. ^44^

We identified a number of genes linked to immune functions that were under selection in our sample. This was reflected in our functional enrichment and pathway enrichment analyses where biological processes and pathways linked to the immune system were strongly implicated. This is expected since immune function related genes are known to be hypervariable by virtue of a need to adapt to new and ever-changing infectious pathogens ^45^.

This concurrent selection from apparently disparate biological processes, raises a number of extremely interesting questions about the interplay between the immune system and neuropsychiatric phenotypes such as psychosis. As an example, the KIR family of genes are implicated in a number of immune related functions, and also influence neurodevelopment and immune reactions in the brain. The *KIR3DL1* has been linked to several aspects of natural killer cell responses, which in turn have been linked to susceptibility to multiple sclerosis, especially in African-Americans ^46^. The KIR2DL4 immunoglobulin like receptor has also been implicated in the development and maintenance of oligodendrocytes ^47^; and also thought to have been positively selected for enabling uterine tolerance for embryonic implantation in humans ^48^. Similarly, the *KIR3DL2* have also been implicated in the interface between immunity genes and brain development, inflammation and responses to damage. They are the receptors for the major histocompatibility complex class I (MHCI) like HLA-F, and protect neurons from astrocyte induced toxicity, as is seen in ALS ^49^. The *IGSF3* gene forms a complex with Tetraspanin *TSAN7*, which is involved in neurodevelopment (many mutations in this gene are linked to X-linked syndromes), and thus mediates a cross-talk between immune mechanisms and development ^50^.

Findings from GWAS and WGS of schizophrenia have also consistently reported signals from the HLA region on Chromosome 6 ^51^. A study which investigated the role of balancing selection using exome data from modern and archaic humans also found an excess of SNPs across species in a gene set associated with the immune system, of which six were located within genes previously associated with schizophrenia ^52^. Another study also suggests shared genetic pathways link white blood cell indices and complex diseases such as schizophrenia, autoimmune, and coronary heart diseases ^53^.

Although we observed the archaic introgression in a single gene (*AHNAK2*) out of the identified set of putative selected genes, we did not find any evidence of archaic introgression under positive selection in any of the genes that were identified using the PBS analysis. These could be related to the fact that exome sequences have a restricted utility when it comes to finding introgressed regions, as the exonic regions are short and spaced, in the context of the whole genome. Additionally, from an evolutionary standpoint, selection tends to happen either upstream or downstream of the genes in areas such as transcription binding sites, rather than in the exonic regions which are well conserved ^54^. Furthermore, exonic regions are generally under negative selection and therefore may not exhibit differences between modern and archaic hominins. Therefore, even if some highly differentiated exonic segment introgressed, it is most likely to have been weeded out by negative selection from the population, as Neanderthals have many more deleterious SNPs due to their low effective population size ^55^. Thus, while it is not impossible to find an introgressed exonic region being selected for, these are rare in the literature ^56^.

A few previous studies looking for selection signatures in south Asian populations using different methodologies have found evidence for positive selection in genes related to lipid metabolism and glucose uptake and have posited a link between the same and the predilection towards development of type 2 diabetes and obesity ^57^ and height in Andaman Island populations ^58^. Our study provides fresh insights into selection in a population from southern India.

In conclusion, our findings show that families with multiple members affected with severe mental illness can be used to detect signatures of selection. Immune related genes showed the greatest evidence of selection in these families. This underscores the contribution of immune mechanisms and infection susceptibility to the genetic architecture of severe mental illnesses such as schizophrenia.

## Methods

### Sequencing and Quality control

As described in our previous study ^59^, whole exome sequencing was carried out on the Illumina Hiseq NGS platform with libraries prepared using Illumina exome kits. Reads were aligned with the reference human genome hg19 using the Burrows-Wheeler algorithm tool (https://academic.oup.com/bioinformatics/article/25/14/1754/225615).

### Study population

The families were recruited as a part of the Accelerator Program for Discovery in Brain Disorders using Stem Cells (ADBS) study, which has been approved by the ethics committee of the National Institute of Mental Health and Neurosciences, Bengaluru, India. The DNA samples with whole-exome sequencing data taken for PBS analysis (N=82) were from 82 families comprising 165 cases with severe mental illness and 33 familial controls; in addition to 10 individuals as population controls.

### Variant Calling

We used bcftools-1.9 ^60^ to do the variant calling for all our samples from the bam files. First, we used bcftools mpileup to create genotype likelihoods. We used a minimum base quality of 20 and a minimum mapping quality of 20 to accept it as a true variant. We also used an adjusted mapping quality of 50 to downgrade reads containing excessive mismatches (as recommended in bcftools for BWA). Additionally, we annotated the file using FORMAT/DP so we had depth information in the vcf file. The output was then piped to bcftools call. We used -m for multi allelic caller. We only used SNPs which were present in the gnomAD vcf file using -T command.

An example of the code is presented here:

bcftools mpileup --ignore-RG -q 20 -Q 20 -C 50 -r <chr> -a FORMAT/DP -f <ucsc.hg19.fasta> <*.bam> | bcftools call -m -T <gnomead.vcf.gz> -O z -o <out.vcf.gz>

### Filtering

After variant calling, we first did a liftover of the vcf file from hg19 to GRCh37 using picard tools. We kept only snps (using --remove-indels flag). We removed any genotype where the coverage was less than 10x using (--minDP 10) and removed any SNPs where we had more than 50% missing genotype data (using --max-missing 0.5). All these commands were done by using vcftools ^61^. We kept only unrelated individuals from the affected multiplex families and disease free (control group) individuals for further analysis.

### PBS calculation

We took the vcf file generated from the previous step and used vcftools to create a frequency file for both unrelated individuals and the control group. We used bcftools query to extract information about the frequencies of gnomAD vcf files. We only extracted AF_afr (alternate allele frequency of African-American/African ancestry individuals), AF_sas (alternate allele frequency of South Asian ancestry individuals), AN_afr ((total number of alleles in samples of African-American/African ancestry individuals) and AN_sas (total number of alleles in samples of South Asian ancestry individuals) from the info columns from gnomAD vcf files. These frequency data were used to calculate PBS (X, SAS, AFR) [where X is our data consisting of diseased unrelated individuals] using in house code with scikit-allele ^62^. We then extracted the top SNPs by their PBS values and tried to find their impact on phenotype.

### Frequency differentiation between cases and controls

As our target population consists of cases, some of the top PBS values can come from regions which might simply be associated with caseness due to sampling bias. To circumvent this problem, we also calculated the allele frequency differences between case and control data set (|Freq _case_-Freq _control_|). Subsequently, SNPs (> 99.9th percentile distribution of PBS) were only considered as potential targets of selection if they had allele frequency difference between cases and controls < 99.9th percentile and any SNPs with allele frequency difference of > 99.9th percentile were dropped. The rationale being that if the PBS value of a SNP is high due to selection instead of the caseness then the allele frequency difference between cases and controls should not vary as much.

### Identification of top candidate genes

We used python to select genes at the top 0.1% (> 99.9th percentile) of the overall PBS distribution and calculated the number of SNPs per gene. We thus identified 75 genes as the putative candidates for selection in the target population.

### Analyses of functional enrichment

To perform the enrichment analysis, we used the set of genes obtained via above method. For GO enrichment, we used the online tool in http://www.geneontology.org/page/go-enrichment-analysis; (last accessed December 8, 2020). We analyzed each of the four gene lists with the three GO categories (biological processes, cellular components, and molecular function) using FDR correction with a significance based on P value < 0.05 (ran on 8th Dec, 2020).

The pathway over-representation analysis on the same gene sets was run using the IMPaLA online tool ^63^ available at: http://impala.molgen.mpg.de (ran on 8th Dec, 2020) and we considered only pathways with a Q-value less than 0.05 to minimize the false positives because Q-value < 0.05 implies only 5% of results can be false positives.

### Detection of archaic introgression in selected gene regions

We applied *haplostrips* tool to detect archaic introgression ^64^ in putative positively selected genes. Haplostrips uses phased genetic data and visualizes polymorphisms of a particular genomic region by clustering and sorting haplotypes independently. The data was phased using shapeit ^65^ with 1000 genome third phase reference ^66^.

## Data Availability

The data is available upon request from the corresponding author.

## Acknowledgements

This research is funded by the Accelerator program for discovery in brain disorders using stem cells (ADBS) (jointly funded by the Department of Biotechnology, Government of India, and the Pratiksha trust; Grant BT/PR17316/MED/31/326/2015). The authors are grateful to all the patients, and their family members who participated in the study.

## Author contributions

Study concept and design – SJ, MP and MM, Acquisition of data – RKN, BV, SJ, Bio-informatics and Data Analysis – AKP, AV, MM, Interpretation of data – JM, SJ, MP, MM, Manuscript drafting – JM, Critical revision of manuscript – All

## Additional Information

The authors declare no competing interests

## Supplementary data

**Supplementary Table S1:** List of genes with 1 SNP per gene considering SNPs that fall within 99.9^th^ percentile of PBS distribution and are out of 99.9^th^ percentile of difference in the frequency between cases and controls (marked with *, otherwise NA); the list also contains frequency of each SNP in cases and controls, and also in continental populations extracted from gnomAD.

**Supplementary Table S2:** List of genes with 2 or more SNPs per gene, considering SNPs that fall within 99.9^th^ percentile of PBS distribution but are out of 99.9^th^ percentile of difference in the frequency between cases and controls (marked with *, otherwise NA); the list contains frequency of each SNP in cases and controls, and also in continental populations extracted from gnomAD.

**Supplementary Table S3:** Final list of multiSNP genes (>2 SNP) and single SNP genes with average PBS values after passing the frequency difference cutoff.

**Supplementary Table S4:** List of multiSNP genes with plausible association to Schizophrenia, Cognitive ability and Intelligence, and Parkinson’s disease (PD) or Alzheimer’s disease (AD), their description and reference sources.

**Supplementary Table S5:** GO for enrichment of MultiSNP genes in biological processes.

**Supplementary Table S6:** GO for enrichment of MultiSNP genes in molecular functions.

**Supplementary Table S7:** GO for enrichment of MultiSNP genes in Cellular components.

**Supplementary Table S8:** IMPaLa pathways enrichment analysis results for the list of 75 multiSNP genes altogether.

## References

1. McGrath, J., Saha, S., Chant, D. & Welham, J. Schizophrenia: A Concise Overview of Incidence, Prevalence, and Mortality. Epidemiol Rev 30, 67–76 (2008).

2. Rowland, T. A. & Marwaha, S. Epidemiology and risk factors for bipolar disorder. Ther Adv Psychopharmacol 8, 251–269 (2018).

3. Wray, N. R. & Visscher, P. M. Narrowing the Boundaries of the Genetic Architecture of Schizophrenia. Schizophr Bull 36, 14–23 (2010).

4. Liu, C., Everall, I., Pantelis, C. & Bousman, C. Interrogating the Evolutionary Paradox of Schizophrenia: A Novel Framework and Evidence Supporting Recent Negative Selection of Schizophrenia Risk Alleles. Front. Genet. 10, (2019).

5. Pattabiraman, K., Muchnik, S. K. & Sestan, N. The evolution of the human brain and disease susceptibility. Curr Opin Genet Dev 65, 91–97 (2020).

6. Guo, J., Yang, J. & Visscher, P. M. Leveraging GWAS for complex traits to detect signatures of natural selection in humans. Curr Opin Genet Dev 53, 9–14 (2018).

7. Polimanti, R. & Gelernter, J. Widespread signatures of positive selection in common risk alleles associated to autism spectrum disorder. PLoS Genet 13, e1006618 (2017).

8. Srinivasan, S. et al. Genetic Markers of Human Evolution Are Enriched in Schizophrenia. Biol. Psychiatry 80, 284–292 (2016).

9. Xiang, B. et al. The role of genes affected by human evolution marker GNA13 in schizophrenia. Prog Neuropsychopharmacol Biol Psychiatry 98, 109764 (2020).

10. Pardiñas, A. F. et al. Common schizophrenia alleles are enriched in mutation-intolerant genes and in regions under strong background selection. Nature Genetics 50, 381–389 (2018).

11. Yao, Y. et al. No Evidence for Widespread Positive Selection Signatures in Common Risk Alleles Associated with Schizophrenia. Schizophr Bull 46, 603–611 (2020).

12. Peng, Y. et al. Down-Regulation of EPAS1 Transcription and Genetic Adaptation of Tibetans to High-Altitude Hypoxia. Mol Biol Evol 34, 818–830 (2017).

13. Zhou, S. et al. Genetic architecture and adaptations of Nunavik Inuit. Proc Natl Acad Sci U S A 116, 16012–16017 (2019).

14. Hallmark, B. et al. Genomic Evidence of Local Adaptation to Climate and Diet in Indigenous Siberians. Mol Biol Evol 36, 315–327 (2019).

15. Ávila-Arcos, M. C. et al. Population History and Gene Divergence in Native Mexicans Inferred from 76 Human Exomes. Molecular Biology and Evolution 37, 994–1006 (2020).

16. Reynolds, A. W. et al. Comparing signals of natural selection between three Indigenous North American populations. PNAS 116, 9312–9317 (2019).

17. Dannemann, M. & Racimo, F. Something old, something borrowed: admixture and adaptation in human evolution. Curr Opin Genet Dev 53, 1–8 (2018).

18. Browning, S. R., Browning, B. L., Zhou, Y., Tucci, S. & Akey, J. M. Analysis of Human Sequence Data Reveals Two Pulses of Archaic Denisovan Admixture. Cell 173, 53-61.e9 (2018).

19. Jacobs, G. S. et al. Multiple Deeply Divergent Denisovan Ancestries in Papuans. Cell 177, 1010-1021.e32 (2019).

20. Zeberg, H. & Pääbo, S. The major genetic risk factor for severe COVID-19 is inherited from Neanderthals. Nature 587, 610–612 (2020).

21. Dolgova, O. & Lao, O. Evolutionary and Medical Consequences of Archaic Introgression into Modern Human Genomes. Genes (Basel) 9, (2018).

22. Gregory, M. D. et al. Neanderthal-Derived Genetic Variation is Associated with Functional Connectivity in the Brains of Living Humans. Brain Connect 11, 38–44 (2021).

23. Narasimhan, V. M. et al. The formation of human populations in South and Central Asia. Science 365, (2019).

24. Pathak, A. K. et al. The Genetic Ancestry of Modern Indus Valley Populations from Northwest India. The American Journal of Human Genetics 103, 918–929 (2018).

25. Viswanath, B. et al. Discovery biology of neuropsychiatric syndromes (DBNS): a center for integrating clinical medicine and basic science. BMC psychiatry 18, 106 (2018).

26. Yi, X. et al. Sequencing of Fifty Human Exomes Reveals Adaptation to High Altitude. Science 329, 75–78 (2010).

27. Wright, S. M. The genetical structure of populations. Ann Eugen 15, 323–354 (1951).

28. Karczewski, K. J. et al. The mutational constraint spectrum quantified from variation in 141,456 humans. Nature 581, 434–443 (2020).

29. Marnetto, D. & Huerta-Sánchez, E. M. Haplostrips: revealing population structure through haplotype visualization. Methods in Ecology and Evolution 8, 1389–1392 (2017).

30. Huang, Y. et al. Recent Adaptive Events in Human Brain Revealed by Meta-Analysis of Positively Selected Genes. PLOS ONE 8, e61280 (2013).

31. Bergen, S. E. et al. Genome-wide association study in a Swedish population yields support for greater CNV and MHC involvement in schizophrenia compared with bipolar disorder. Molecular Psychiatry 17, 880–886 (2012).

32. Ripke, S. et al. Genome-wide association analysis identifies 13 new risk loci for schizophrenia. Nature Genetics 45, 1150–1159 (2013).

33. Ripke, S. et al. Biological insights from 108 schizophrenia-associated genetic loci. Nature 511, 421–427 (2014).

34. Bacchelli, E. et al. An integrated analysis of rare CNV and exome variation in Autism Spectrum Disorder using the Infinium PsychArray. Scientific Reports 10, 3198 (2020).

35. Anney, R. J. L. et al. Conduct disorder and ADHD: Evaluation of conduct problems as a categorical and quantitative trait in the international multicentre ADHD genetics study. American Journal of Medical Genetics Part B: Neuropsychiatric Genetics 147B, 1369–1378 (2008).

36. Hill, W. D. et al. A combined analysis of genetically correlated traits identifies 187 loci and a role for neurogenesis and myelination in intelligence. Molecular Psychiatry 24, 169–181 (2019).

37. Lam, M. et al. Pleiotropic Meta-Analysis of Cognition, Education, and Schizophrenia Differentiates Roles of Early Neurodevelopmental and Adult Synaptic Pathways. The American Journal of Human Genetics 105, 334–350 (2019).

38. Rietveld, C. A. et al. Common genetic variants associated with cognitive performance identified using the proxy-phenotype method. PNAS 111, 13790–13794 (2014).

39. Nicolas, A. et al. Genome-wide Analyses Identify KIF5A as a Novel ALS Gene. Neuron 97, 1268-1283.e6 (2018).

40. Simón-Sánchez, J. et al. Genome-wide association study reveals genetic risk underlying Parkinson’s disease. Nature Genetics 41, 1308–1312 (2009).

41. Figley, M. D. & DiAntonio, A. The SARM1 axon degeneration pathway: control of the NAD+ metabolome regulates axon survival in health and disease. Curr Opin Neurobiol 63, 59–66 (2020).

42. Fang, L. et al. Comparative analyses of sperm DNA methylomes among human, mouse and cattle provide insights into epigenomic evolution and complex traits. Epigenetics 14, 260–276 (2019).

43. Hart, P. H., Norval, M., Byrne, S. N. & Rhodes, L. E. Exposure to Ultraviolet Radiation in the Modulation of Human Diseases. Annu Rev Pathol 14, 55–81 (2019).

44. Shen, Y., Ha, W., Zeng, W., Queen, D. & Liu, L. Exome sequencing identifies novel mutation signatures of UV radiation and trichostatin A in primary human keratinocytes. Scientific Reports 10, 4943 (2020).

45. Shultz, A. J. & Sackton, T. B. Immune genes are hotspots of shared positive selection across birds and mammals. eLife 8, e41815 (2019).

46. Hollenbach, J. A., Pando, M. J., Caillier, S. J., Gourraud, P.-A. & Oksenberg, J. R. The killer immunoglobulin-like receptor KIR3DL1 in combination with HLA-Bw4 is protective against multiple sclerosis in African Americans. Genes & Immunity 17, 199–202 (2016).

47. Banerjee, P. P. et al. KIR2DL4-HLAG interaction at human NK cell-oligodendrocyte interfaces regulates IFN-γ-mediated effects. Molecular Immunology 115, 39–55 (2019).

48. Vallender, E. J. Chapter 1 - Genetics of human brain evolution. in Progress in Brain Research (ed. Hofman, M. A.) vol. 250 3–39 (Elsevier, 2019).

49. Song, S. et al. Major histocompatibility complex class I molecules protect motor neurons from astrocyte-induced toxicity in amyotrophic lateral sclerosis. Nature Medicine 22, 397–403 (2016).

50. Perot, B. P. & Ménager, M. M. Tetraspanin 7 and its closest paralog tetraspanin 6: membrane organizers with key functions in brain development, viral infection, innate immunity, diabetes and cancer. Med Microbiol Immunol 209, 427–436 (2020).

51. Birnbaum, R. & Weinberger, D. R. A Genetics Perspective on the Role of the (Neuro)Immune System in Schizophrenia. Schizophrenia Research 217, 105–113 (2020).

52. Viscardi, L. H. et al. Searching for ancient balanced polymorphisms shared between Neanderthals and Modern Humans. Genet Mol Biol 41, 67–81 (2018).

53. Astle, W. J. et al. The Allelic Landscape of Human Blood Cell Trait Variation and Links to Common Complex Disease. Cell 167, 1415-1429.e19 (2016).

54. Dannemann, M., Prüfer, K. & Kelso, J. Functional implications of Neandertal introgression in modern humans. Genome Biology 18, 61 (2017).

55. Juric, I., Aeschbacher, S. & Coop, G. The Strength of Selection against Neanderthal Introgression. PLOS Genetics 12, e1006340 (2016).

56. Racimo, F., Sankararaman, S., Nielsen, R. & Huerta-Sánchez, E. Evidence for archaic adaptive introgression in humans. Nat Rev Genet 16, 359–371 (2015).

57. Metspalu, M. et al. Shared and unique components of human population structure and genome-wide signals of positive selection in South Asia. Am J Hum Genet 89, 731–744 (2011).

58. Mondal, M. et al. Genomic analysis of Andamanese provides insights into ancient human migration into Asia and adaptation. Nat Genet 48, 1066–1070 (2016).

59. Ganesh, S. et al. Exome sequencing in families with severe mental illness identifies novel and rare variants in genes implicated in Mendelian neuropsychiatric syndromes. Psychiatry and Clinical Neurosciences 73, 11–19 (2019).

60. Li, H. A statistical framework for SNP calling, mutation discovery, association mapping and population genetical parameter estimation from sequencing data. Bioinformatics 27, 2987–2993 (2011).

61. Danecek, P. et al. The variant call format and VCFtools. Bioinformatics 27, 2156–2158 (2011).

62. Miles, A. & Harding, N. cggh/scikit-allel: v1. 1.8 (Version v1. 1.8). Zenodo. Available at: doi 10, (2017).

63. Kamburov, A., Cavill, R., Ebbels, T. M. D., Herwig, R. & Keun, H. C. Integrated pathway-level analysis of transcriptomics and metabolomics data with IMPaLA. Bioinformatics 27, 2917–2918 (2011).

64. Green, R. E. et al. A Draft Sequence of the Neandertal Genome. Science 328, 710–722 (2010).

65. Delaneau, O. & Marchini, J. Integrating sequence and array data to create an improved 1000 Genomes Project haplotype reference panel. Nature Communications 5, 3934 (2014).

66. Auton, A. et al. A global reference for human genetic variation. Nature 526, 68–74 (2015).

